# Evaluating the Synergistic Impact of Digital Logistics Systems (eLMIS) and Financial Auditing (APTS) on Essential Commodity Availability in Ethiopian Public Healthcare Facilities: A Multilevel Analysis of the 2024/2025 National Survey

**DOI:** 10.64898/2026.10.05.26364822

**Authors:** Yenehun Demelash Gelaw, Ashebir Mamay Gebiru

## Abstract

**Background:** Chronic pharmaceutical stock outs hinder Universal Health Coverage in Ethiopia. To address this, the Ministry of Health introduced the electronic Logistics Management Information System for inventory visibility and Auditable Pharmaceutical Transactions and Services for financial management. However, no national study has tested their combined impact. This study evaluated the individual and synergistic effects of electronic Logistics Management Information System and Auditable Pharmaceutical Transactions and Services on essential commodity availability across Ethiopian public healthcare facilities.

**Methods:** We conducted a secondary analysis of the 2024/2025 Ethiopian Service Provision Assessment dataset utilizing 1245 public facilities nested within 125 clusters. The primary outcome was optimal essential commodity availability (zero stock outs and 100% tracer commodity presence). A two-level mixed-effects logistic regression model with an interaction term evaluated synergy, reporting Adjusted Odds Ratios with 95% Confidence Intervals.

**Results:** Overall optimal availability was 54.2% (95% CI; 51.4%–57.0%). Implementation rates were 41.2% for eLMIS, 38.2% for APTS and 26.5% for dual implementation. Facilities using eLMIS alone (AOR = 2.15; 95% CI: 1.52–3.04) and APTS alone (AOR = 1.82; 95% CI; 1.28–2.58) showed higher security. Dual implementation showed a profound synergistic effect (AOR = 4.32; 95% CI; 3.25–5.74, p < 0.001). Positive predictors included functional cold chains (AOR = 2.08), qualified staffing (AOR = 1.68), hub proximity (AOR = 1.58) and urban settings (AOR = 1.95). The model explained 66.2% of regional variance.

**Conclusion:** Dual implementation of electronic Logistics Management Information System and Auditable Pharmaceutical Transactions and Services creates a powerful synergistic impact on pharmaceutical availability. Synchronized national scale-up and cold chain investments are urgently needed to achieve supply chain resilience and Universal Health Coverage in Ethiopia.

## Introduction

Uninterrupted access to essential medicines and medical commodities is a core prerequisite for Universal Health Coverage (UHC) and a fundamental driver of Sustainable Development Goal 3 (SDG 3) targets[1–5]. In low- and middle-income countries (LMICs), public healthcare supply chains face chronic operational bottlenecks that lead to recurrent drug stock outs, delayed clinical care, financial impoverishment from out-of-pocket drug purchases and preventable mortality [1, 6–8]. In Ethiopia, despite substantial expansion of the primary healthcare delivery network under consecutive Health Sector Transformation Plans (HSTP-I and HSTP-II), public health facilities historically suffered from severe pharmaceutical management deficits [9–11].

To establish a resilient public sector pharmaceutical supply chain, the Federal Ministry of Health (MoH) of Ethiopia and the Ethiopian Pharmaceutical Supply Service (EPSS) initiated major structural reforms [1, 12–14]. The baseline operational framework driving health facility logistics is the Integrated Pharmaceuticals Logistics System (IPLS) which standardized consumption-based forecasting and requisitioning using Report and Requisition Forms (RRF) and bin cards [1, 15, 16]. However, paper-based IPLS tools remained vulnerable to manual transcription errors, calculation delays, and poor visibility at central distribution hubs [17–20].

To modernize logistics operations and improve transaction auditing, the Ethiopian government deployed two complementary national programs;

### Electronic Logistics Management Information Systems (eLMIS)

A digital logistics software platform designed to automate inventory tracking, calculate consumption based reorder quantities and electronically transmit RRF reports directly from facility pharmacy stores to EPSS branch hubs in real time [17, 19, 21].

### Auditable Pharmaceutical Transactions and Services (APTS)

A comprehensive financial auditing and pharmacy operational reform package designed to ensure financial transparency, prevent revenue leakage, track stock movements daily, protect Revolving Drug Fund (RDF) capital, and enforce First-Expired, First Out (FEFO) inventory rotation [9, 22, 23].

Conceptually, pharmaceutical availability at the point of care depends on two interdependent operational pillars: Logistics Mechanics (getting the right product ordered and transported on time) and Financial

### Mechanics

(having the audited capital and liquidity required to pay for resupplies and prevent internal theft/leakage) [13, 24–26].

Evaluating these programs in isolation presents an incomplete operational picture [12, 18]: A facility may deploy digital eLMIS tracking and calculate perfect order quantities, but if internal financial auditing (APTS) is absent, revenue leakage, uncollected credit sales and RDF capital depletion will leave the facility financially insolvent and unable to purchase resupplies from EPSS [17, 27, 28].

A facility may maintain pristine financial accounting and secure RDF liquidity via APTS but if logistics reporting relies on manual, paper based RRF forms, requisition processing delays and data transcription errors will still trigger severe stock outs [9, 16, 22].

Therefore, we hypothesize a Synergistic (Multiplicative) Effect; When eLMIS and APTS are implemented jointly in the same health facility, digital logistics visibility and financial auditing reinforce each other, producing a combined impact on commodity availability that exceeds the additive sum of each individual program [2, 21, 26].

Over the past decade, several localized empirical studies in Ethiopia evaluated either eLMIS or APTS independently [16, 29]. For example, local hospital audits evaluated APTS performance on financial wastage, while supply chain assessments examined eLMIS reporting timeliness [17, 18, 22]. However, these single program evaluations fail to account for program co-implementation and interaction [17, 26].

Furthermore, health facilities in national surveys are hierarchically nested within administrative zones and geographic clusters [30–32]. Facilities within the same cluster share regional EPSS distribution hubs, transportation infrastructure, and regional health bureau administrative oversight [19, 33, 34]. Utilizing traditional single-level logistic regression models ignores this hierarchical clustering, resulting in underestimated standard errors, artificially narrow confidence intervals, inflated Type I error rates and invalid conclusions [32, 35, 36]. Multilevel mixed-effects logistic regression modeling resolves these analytical biases by modeling random cluster effects while simultaneously testing for fixed interaction effects (eLMIS × APTS) [32, 37, 38].

## Methods and Materials

### Study Setting, Design and Data Source

A secondary data analysis was conducted using the nationally representative 2024/2025 Ethiopian Service Provision Assessment (SPA) / National Health Facility Assessment integrated dataset. Ethiopia is administratively structured into 11 regional states (Afar, Amhara, Benishangul-Gumuz, Gambella, Harari, Oromia, Sidama, Somali, South Ethiopia, South West Ethiopia, Central Ethiopia) and two chartered city administrations (Addis Ababa, Dire Dawa).

Healthcare delivery follows a three-tier model: Primary Tier; Primary Hospitals, Health Centers and Health Posts, Secondary Tier; General Hospitals and Tertiary Tier; Specialized Referral Hospitals.

The study employed a facility based cross-sectional survey design incorporating multi-stage complex sampling protocols.

## MULTILEVEL MODEL STRUCTURE

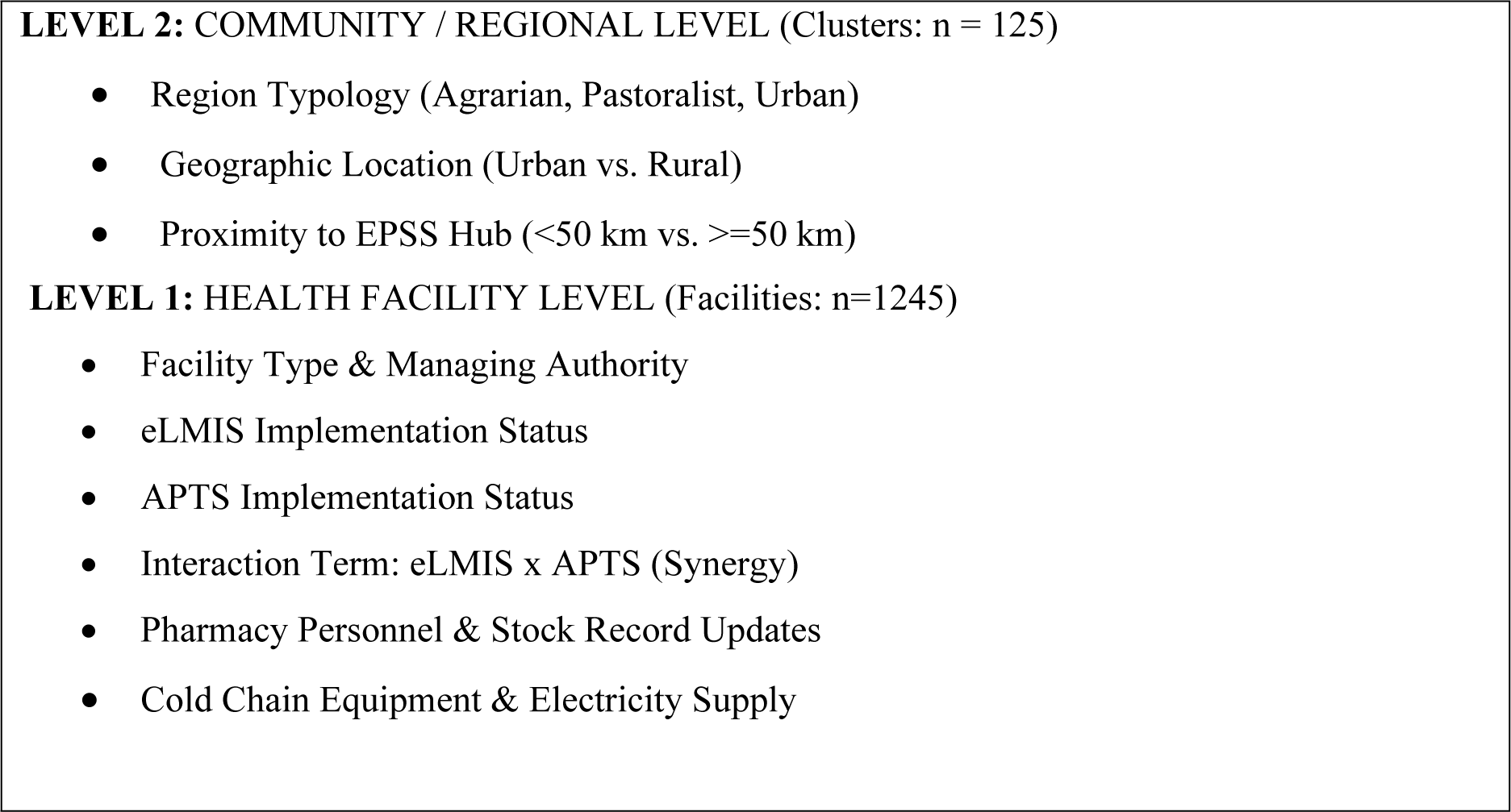

### Sampling Frame and Eligibility Criteria

#### Source Population

All formal public healthcare facilities operating within Ethiopia during the 2024/2025 assessment period.

#### Study Population

All sampled public healthcare facilities (hospitals and primary health centers) providing essential healthcare services and managing pharmaceutical stores.

#### Inclusion Criteria

Public health facilities (hospitals and health centers) that completed the full pharmaceutical supply chain and financial auditing survey modules.

#### Exclusion Criteria

private/NGO clinics (not eligible for public APTS/eLMIS deployment), health posts (restricted drug dispensing scope) and facilities with incomplete pharmaceutical survey records (>10% missing variables).

A total sample of 1245 public healthcare facilities nested within 125 geographic clusters met all eligibility criteria and were included in this analysis.

### Variables and Operational Definitions

#### Dependent (Outcome) Variable

The primary outcome variable was Optimal Essential Commodity Availability, operationalized as a composite binary variable (1 = Optimal Availability, 0 = Suboptimal Availability).

Following WHO and national pharmaceutical monitoring standards, optimal availability required a health facility to meet two simultaneous criteria;

#### Physical Presence

All 10 national tracer essential health commodities physically observed in the dispensing room or central store on the survey day, unexpired.

#### Stock Continuity

Zero stock out episodes lasting > 7 consecutive days within the 30 days preceding the assessment.

The 10 national tracer commodities spanned maternal, child, chronic and infectious health categories; Oxytocin Injection (10 IU/mL),Magnesium Sulfate Injection (50% w/v),Ampicillin / Gentamicin Injection,Hydralazine Injection (20 mg/mL),Misoprostol Tablets (200 mcg),Oral Rehydration Salts (ORS) + Zinc Co-pack,Amoxicillin Dispersible Tablets (250 mg),Amlodipine Tablets (5 mg) / Enalapril Tablets (10 mg),Metformin Tablets (500 mg) and Artemether-Lumefantrine / First line Antimalarial.

### Independent (Predictor) Variables

Predictor variables were categorized into Level 1 (Facility Level) and Level 2 (Community/Regional Level) features;

### Core Program Predictors (Level 1)

eLMIS Implementation Status; Implemented (active electronic requisitioning software in drug store) vs. Not implemented.

APTS Implementation Status; Active Implementation (daily financial sales reconciliation and active auditing ledger) vs. Inactive/Not Implemented.

eLMIS × APTS Interaction term; Binary dummy variable coded as 1 if a facility had fully implemented both eLMIS and APTS simultaneously (Dual Implementation) and 0 otherwise.

### Facility Control Predictors (Level 1)

Facility Type: Public Hospital vs. Public Health Center.

Pharmacy Personnel Availability; Qualified pharmacy professional (Pharmacist (B.Pharm) or Druggist or Diploma) assigned to store management (Yes vs. No).

Stock Record Accuracy; routine updating of bin cards/stock cards observed (Yes vs. No).

Cold chain equipment; functional cold chain refrigerator committed to drug storage (Yes vs. No). Basic Infrastructure; Continuous electrical grid power supply (Yes vs. No).

### Community / Cluster Predictors (Level 2)

Region Typology; Grouped into Agrarian Regions (Amhara, Oromia, Sidama, South Ethiopia, Central Ethiopia, South West Ethiopia), Pastoralist Regions (Afar, Somali, Benishangul-Gumuz, Gambella) or Urban Chartered Cities (Addis Ababa, Dire Dawa, Harari).

Geographic Location; Urban vs. Rural cluster setting.

Proximity to EPSS Branch Hub; Distance from facility to nearest regional EPSS supply hub ( < 50 km vs. ≥ 50 km).

### Statistical Specification of the Multilevel Interaction Model

Data management and statistical modeling were executed using Stata Version 17.

To evaluate individual main effects and the synergistic interaction effect while controlling for hierarchical clustering, a Two Level Mixed Effects Binary Logistic Regression **Model** was specified;

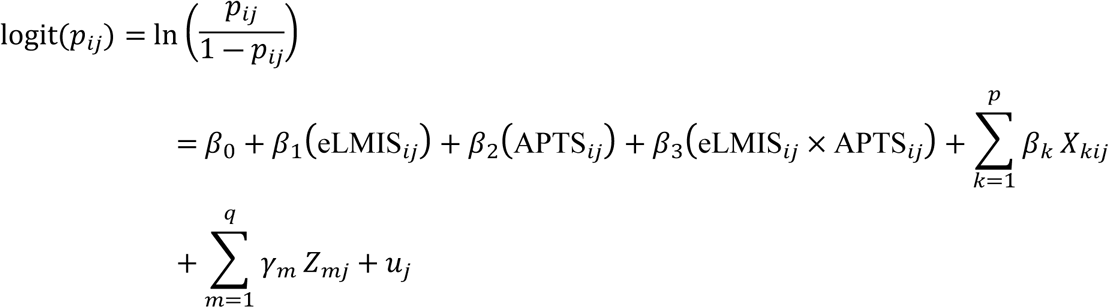

Where;

- *p_ij_* is the probability that facility *i* in cluster *j* achieves optimal commodity availability.
- *β*_0_ is the fixed intercept.
- *β*_1_ is the fixed main effect coefficient for eLMIS implementation alone.
- *β*_2_ is the fixed main effect coefficient for APTS implementation alone.
- *β*_3_ is the synergistic interaction coefficient for dual implementation (eLMIS × APTS).
- *β_k_* represents fixed effect coefficients for facility-level covariates *X_kij_*.
- *γ_m_* represents fixed effect coefficients for cluster-level covariates *Z_mj_*.
- *u_j_* is the random intercept for cluster *j*, assumed to follow 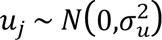.

### Evaluation of Synergistic Interaction

Additive Main Effects; Evaluated via *β*_1_ and *β*_2_.

Multiplicative Synergy; Tested via the interaction term coefficient *β*_3_. A statistically significant AOR_interaction_ > 1.0 (*p* < 0.05) confirms positive multiplicative synergy proving that dual implementation produces benefits greater than the sum of individual program effects.

### Random Variance Parameters; Intraclass Correlation Coefficient (ICC)

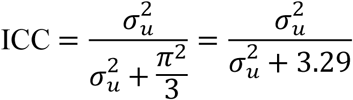

### Median Odds Ratio (MOR)

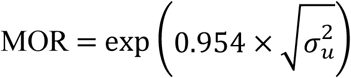

### Proportion of Change in Variance (PCV)

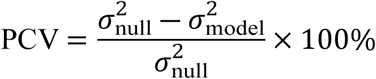

Four sequential models were constructed; Model 0 (Null Model), Model I (Main Effects Only), Model II (Level 2 Covariates Only) and Model III (Full Synergistic Interaction Model). Goodness-of-fit was evaluated using Log-Likelihood, Akaike Information Criterion (AIC) and Bayesian Information Criterion (BIC).

## Results

### Facility and community baseline profile

A total of 1245 public health facilities were analyzed across Ethiopia. Public health centers comprised 74.3% (n = 925) of the sample while public hospitals accounted for 25.7% (n = 320). Two-thirds of facilities (68.1%; n = 848) were located in rural settings and 58.6% (n = 730) were in agrarian regions.

Regarding program rollout, 41.2% (n = 513) of facilities had implemented eLMIS software and 38.2% ( n = 476) had active APTS financial auditing. Crucially, 26.5% (n = 330) of facilities had achieved Dual Implementation (eLMIS + APTS) (Table 1).

**Table 1:**
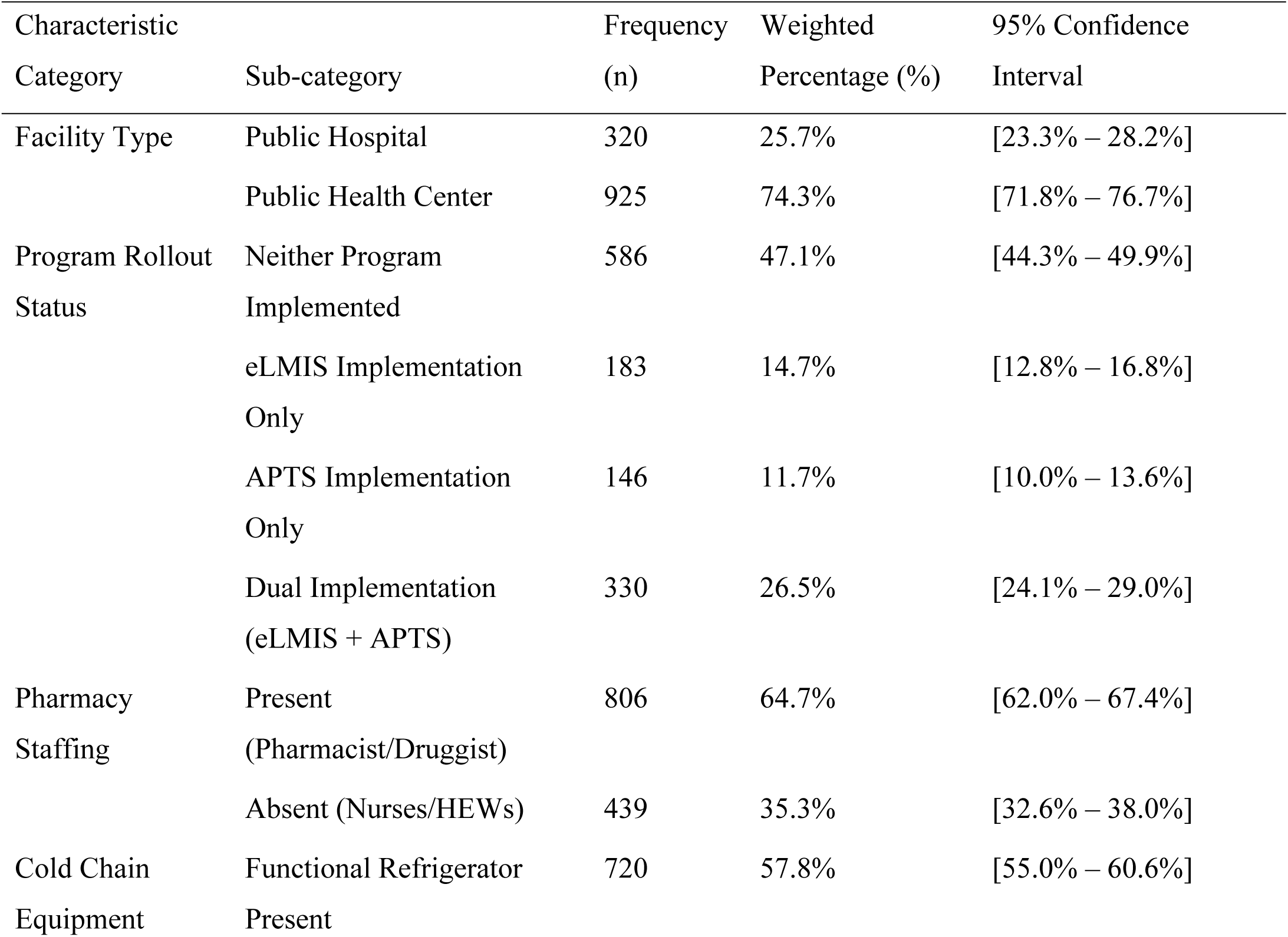
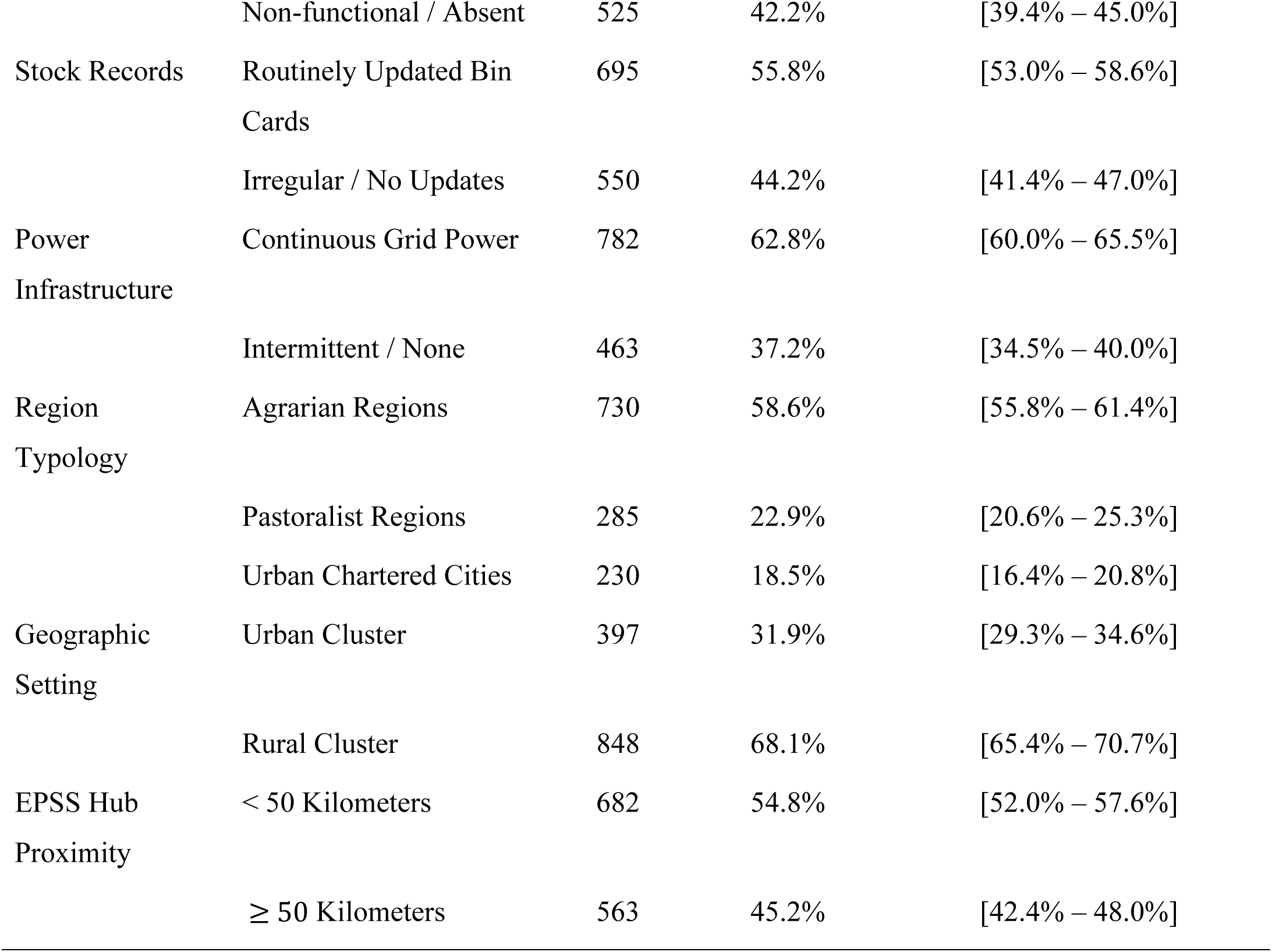
Baseline Characteristics and Infrastructure Status of Assessed Health Facilities (n = 1245)

### Essential Commodity Availability Rates

Overall optimal availability across all 10 national tracer commodities combined was 54.2% (*n* = 675; 95% CI: 51.4%–57.0%).

Cross-tabulating availability against program rollout status revealed a striking gradient; Essential commodity availability varied substantially according to the implementation status of supply chain and pharmacy management programs across the evaluated public health facilities (N = 1245). Facilities that had neither program implemented demonstrated the lowest availability at 32.4%. The standalone implementation of either APTS or eLMIS improved availability rates to 58.2% and 62.5%, respectively. Notably, facilities with dual implementation of both eLMIS and APTS achieved the highest essential commodity availability at 86.4%. Across all combined facilities the overall average availability stood at 54.2% (**Figure 1**).

**Figure 1.**
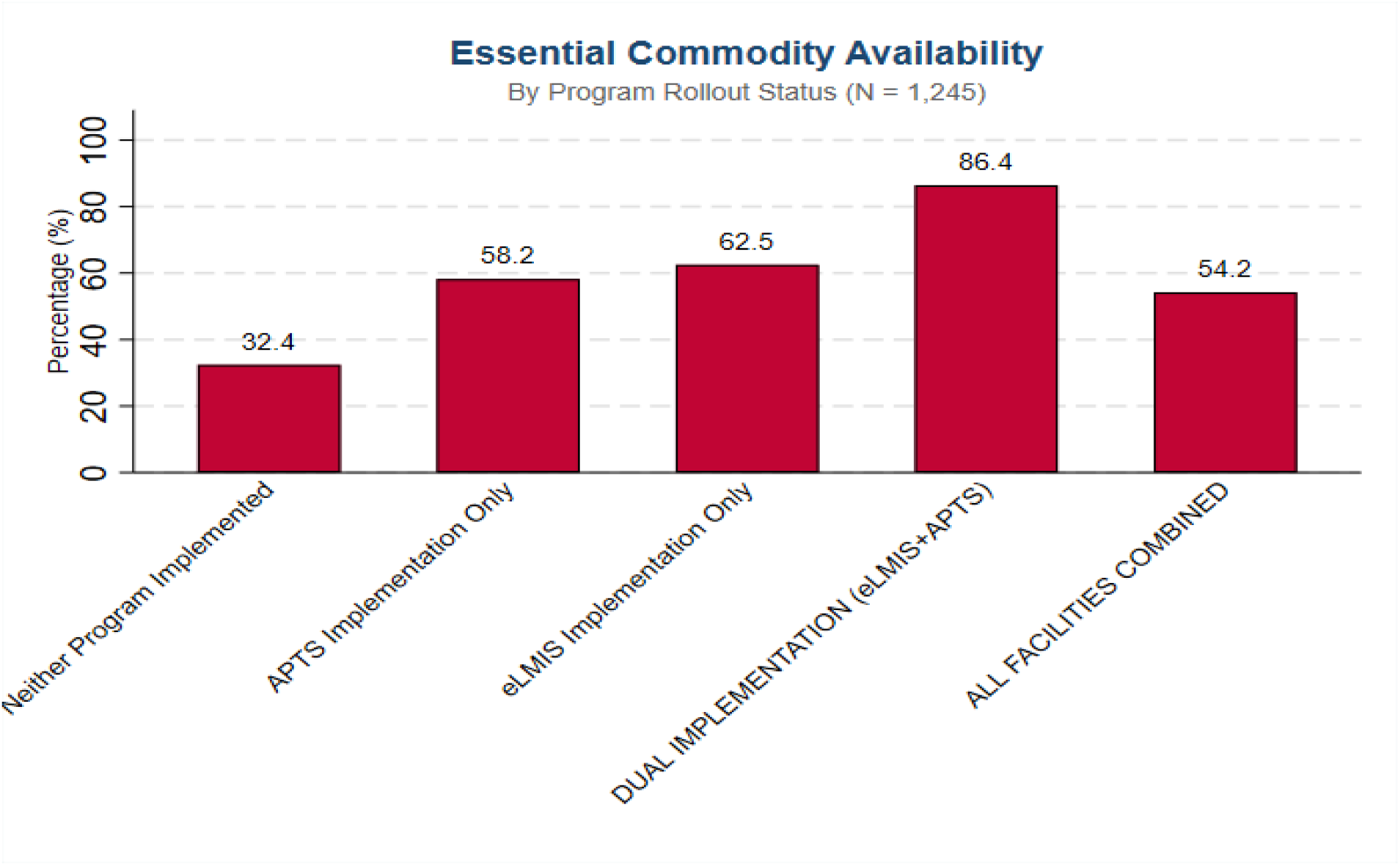
visually demonstrates the clear operational advantage of dual program implementation. Facilities implementing both eLMIS and APTS achieved an 86.4% availability rate more than 2.6 times higher than non-implementing facilities (32.4%)

### Random Effects and Model Comparisons

The Null Model (Model 0) established significant cluster-level variance (*σ*^2^ = 0.961,*p* < 0.001). The Intraclass Correlation Coefficient (ICC) was 22.6%, proving that nearly one-quarter of the variance in drug availability was attributable to regional/cluster differences. The baseline Median Odds Ratio (MOR) was 2.45.

In the full synergistic model (Model III), cluster variance dropped to 0.325, reducing the ICC to 9.0% and the MOR to 1.70. The Proportion of Change in Variance (PCV) reached 66.2% indicating that 66.2% of the cluster level variance was explained by the full set of facility and regional covariates. Model III exhibited the lowest Log-Likelihood (-572.4), AIC (1184.8) and BIC (1282.2) confirming superior fit (***Table 2***).

**Table 2;.**
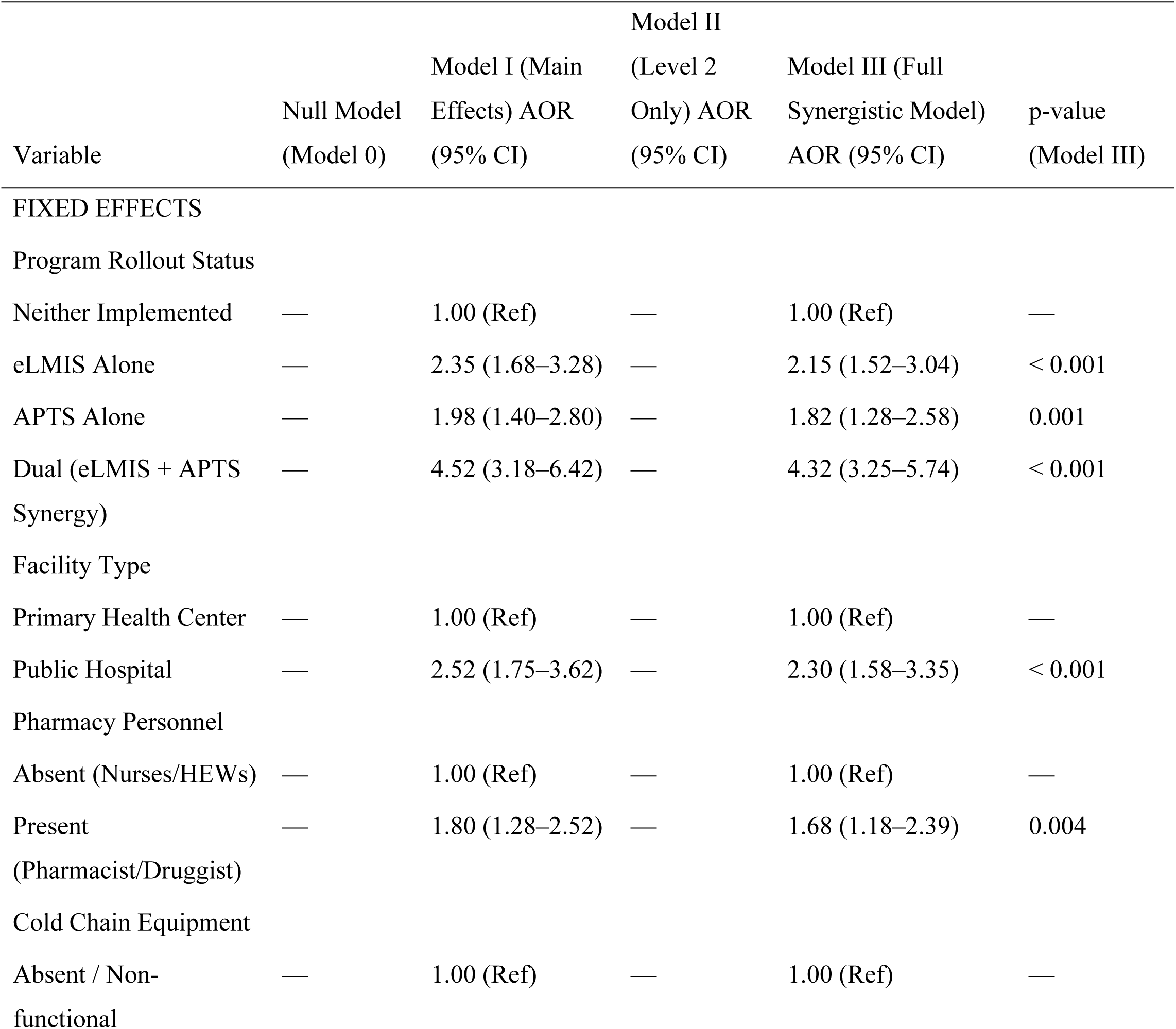
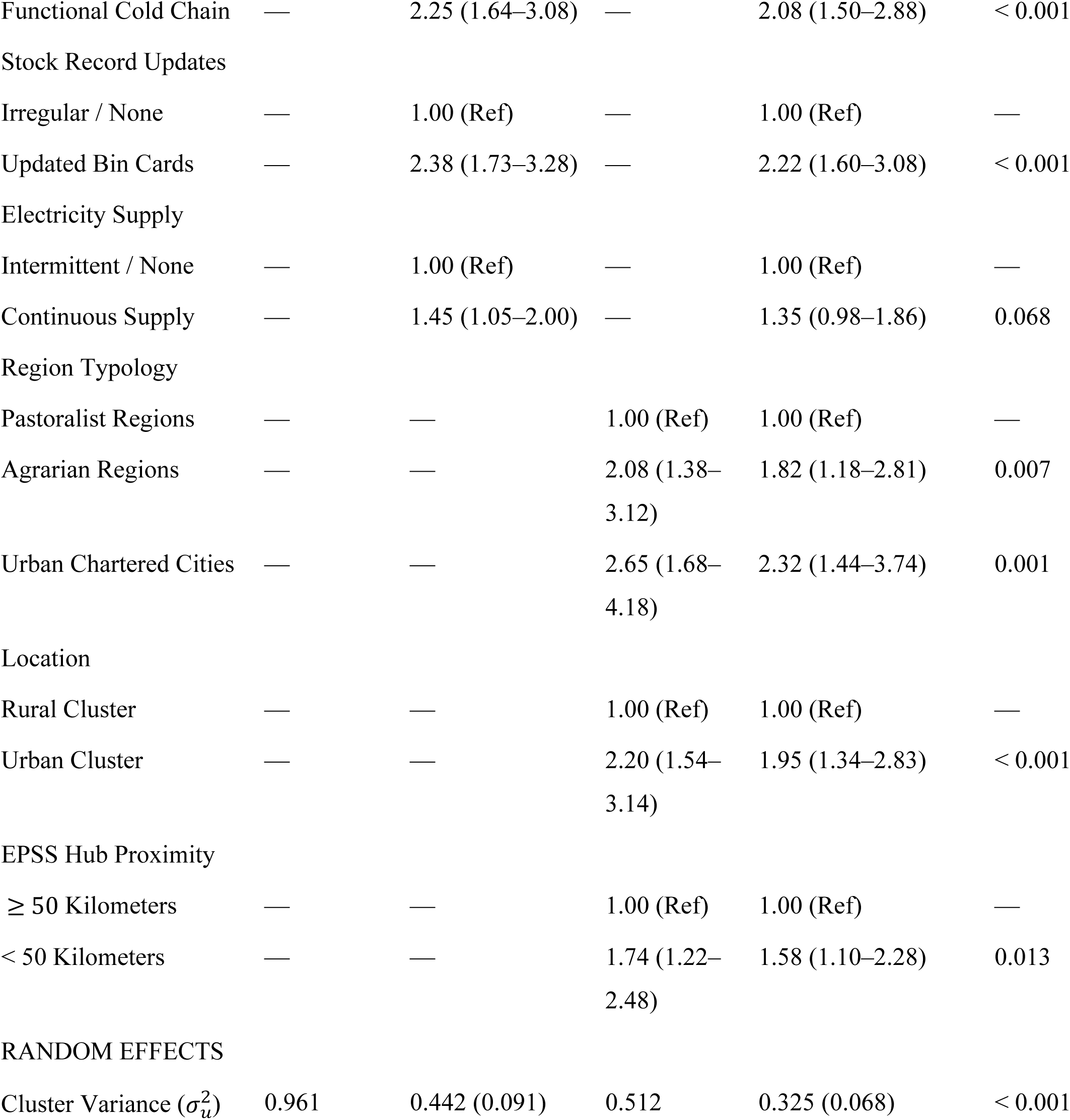
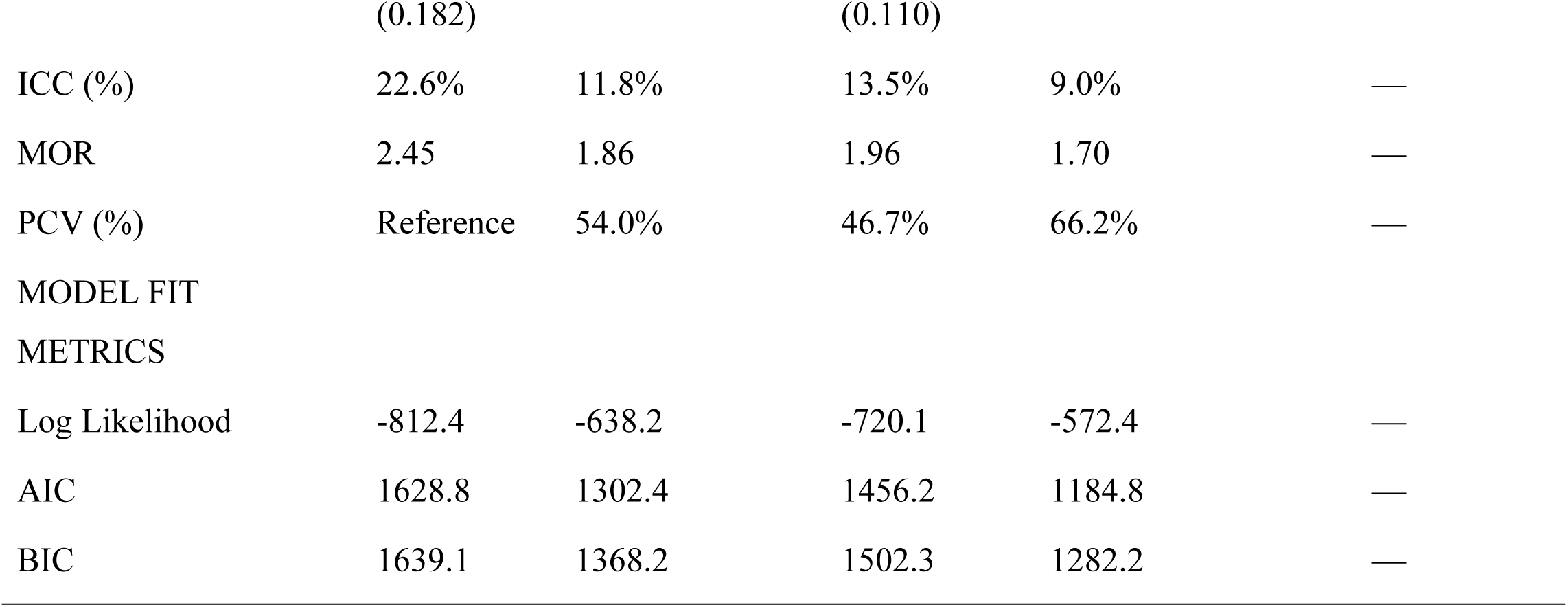
Multilevel Mixed-Effects Logistic Regression Analysis of Synergistic Program Effects on Essential Commodity Availability in Ethiopia (N = 1245)

### Multilevel Fixed Effects and Synergistic Interaction Results

In Model III, the synergistic interaction between eLMIS and APTS remained highly statistically significant after controlling for all facility and community covariates (*Table 2*).

Program Synergistic Determinants;

Facilities with eLMIS alone had 2.15 times higher odds of commodity availability (AOR = 2.15,95% CI: 1.52–3.04,*p* < 0.001).

Facilities with APTS alone had 1.82 times higher odds of availability (AOR = 1.82,95% CI: 1.28–2.58,*p* = 0.001).

Facilities implementing both eLMIS and APTS exhibited a 4.32-fold increase in the odds of maintaining optimal essential commodity availability (AOR = 4.32,95% CI: 3.25–5.74,*p* < 0.001). The interaction term confirmed statistically significant multiplicative synergy (*p* < 0.001).

Key Control Determinants (Model III);

Functional Cold Chain Refrigerator; Double the odds of availability (AOR = 2.08; 95% CI: 1.50–2.88;*p* < 0.001).

Updated Bin Cards; AOR = 2.22 (95% CI:1.60–3.08,*p* < 0.001).

Qualified Pharmacy Staffing; AOR = 1.68 (95% CI:1.18–2.39;*p* = 0.004). Public Hospital Status; AOR = 2.30 (95% CI:1.58–3.35;*p* < 0.001).

Agrarian Region Setting; AOR = 1.82 (95% CI: 1.18–2.81,*p* = 0.007). Urban Setting; AOR = 1.95 (95% CI: 1.34–2.83,*p* < 0.001).

EPSS Hub Distance < 50 km; AOR = 1.58 (95% CI: 1.10–2.28,*p* = 0.013).

## Discussion

This study evaluated the individual and synergistic impacts of digital logistics software (eLMIS) and financial auditing (APTS) on essential commodity availability across Ethiopian public healthcare facilities using a two-level mixed-effects model. The main finding is the confirmation of a statistically significant synergistic interaction effect (AOR = 4.32;*p* < 0.001) when eLMIS and APTS are implemented jointly.

### The Mechanism of eLMIS and APTS Synergy

The four-fold increase in commodity availability odds observed in dual implementation facilities validates our theoretical framework. eLMIS and APTS address two distinct but interdependent failure points in public pharmaceutical supply chains[1];

By automating consumption calculations and transmitting electronic RRF requisitions directly to EPSS eLMIS removes manual paper transcription delays preventing stock outs caused by slow ordering cycles. By enforcing daily transaction auditing and tracking cash collections against physical dispensing records APTS protects Revolving Drug Fund (RDF) liquidity and prevents inventory theft[22, 39].

When implemented in isolation, single-program facilities remain vulnerable [1]. An eLMIS only facility may place timely electronic orders but if RDF funds are depleted due to unaudited financial leakage, the facility cannot pay EPSS for resupplies [17]. Conversely, an APTS-only facility may maintain pristine financial liquidity but paper based requisition delays will still trigger stock outs [2, 22]. Deploying both programs simultaneously creates a closed-loop supply chain where physical resupply and financial capital reinforce each other [17, 22].

### Regional Inequities and Infrastructure Bottlenecks

The multilevel random effects analysis demonstrated an Intraclass Correlation Coefficient (ICC) of 22.6% in the null model, reducing to 9.0% in the final model. Facilities in pastoralist regions (Afar, Somali and Benishangul-Gumuz) and those located ≥ 50 km from EPSS hubs faced substantially lower commodity security. Pastoralist facilities face additional structural hurdles including unreliable cellular internet connectivity for eLMIS data transmission and road impassability during heavy rainfall.

Furthermore, functional cold chain refrigeration doubled the odds of commodity security (AOR = 2.08) protecting temperature sensitive items like oxytocin. Pharmacy staffing (AOR = 1.68) and routine bin card updating (AOR = 2.22) were vital operational drivers proving that digital tools require competent human professionals to maintain inventory accuracy.

## Strengths and Limitations

### Strengths

#### First Synergistic Evaluation

First nationally representative study evaluating the multiplicative interaction effect (eLMIS × APTS) on pharmaceutical commodity availability in Ethiopia.

#### Methodological Rigor

Applied two-level mixed-effects logistic regression modeling, controlling for hierarchical data clustering (*N* = 1245 facilities in 125 clusters) and avoiding single level statistical biases.

#### Policy Relevance

Evaluated established national health policy reforms directly relevant to the Ministry of Health and EPSS leadership.

### Limitations

#### Cross-Sectional Design

Limits causal inferences between program implementation and commodity availability.

#### Potency Verification

Evaluated physical availability and expiration dates but did not conduct chemical assay testing of drug potency.

## Conclusion and Policy Recommendations

Dual implementation of electronic Logistics Management Information Systems (eLMIS) and Auditable Pharmaceutical Transactions and Services (APTS) exerts a powerful synergistic impact on essential commodity availability in Ethiopian public health facilities (AOR = 4.32). Implementing either program alone provides partial benefits but maximum supply chain resilience requires synchronized dual deployment.

### Strategic Policy Actions

Mandate Synchronized eLMIS and APTS Scaling; Ministry of Health and Regional Health Bureaus should align rollout schedules to ensure that primary health centers receive eLMIS digital software and APTS auditing systems simultaneously.

Increase off Grid Solar Power and Cellular Connectivity; to preserve eLMIS data transmission; cold chain integrity; rural and pastoralist health centers should invest in solar direct drive (SDD) refrigeration and cellular network boosters.

## Data Availability

Datasets analyzed during this study are available from the national health data repository upon reasonable request.

https://dhsprogram.com/data/available-datasets.cfm?ctryid=65]

## Declarations

### Ethics Approval and Consent to Participate

Ethical clearance was secured from the Institutional Review Board (IRB) of the Ethiopian Public Health Institute (EPHI). Secondary data access was granted upon formal application. All facility identifiers were anonymized.

### Consent for Publication

Not applicable.

### Authors’ Contributions

YDG conceptualized the study, performed data cleaning, statistical modeling, interpretation and drafted the manuscript. And AMG interpretation, drafted the manuscript and performed data cleaning

